# The burden and determinants of self-reported non-communicable disease comorbidities among HIV positive adults in Tanzania: Insights from the Tanzania HIV Impact Survey 2022-2023

**DOI:** 10.1101/2025.10.27.25338926

**Authors:** Charles B. Kafaiya, Hillary R. Sebukoto, Angelina M. Lutambi, Gibson Kagaruki, Johnson J. Mshiu, Paul M. Hayuma, Gerald P. Mwing’a, Irene R. Mremi, Jonathan M. Mshana, Prince Mutalemwa, Sia E. Malekia, Kunda J. Steven, Mary T. Mayige

## Abstract

People living with HIV (PLHIV) face an increased risk of developing comorbidities involving HIV and non-communicable diseases (NCDs) caused by a mix of modifiable and non-modifiable risk factors. This study aimed to quantify the burden of NCD-HIV comorbidities and examine their determinants among HIV-positive adults aged ≥ 15 years using data from the 2022– 2023 Tanzania HIV Impact Survey (THIS). The 2022–2023 THIS enrolled 33,663 individuals who underwent HIV testing, of whom 1,850 tested positive. Key variables, including socioeconomic, demographic, and behavioral risk factors and clinical characteristics, were extracted from the main survey database. These variables were modelled as potential determinants of HIV–NCD comorbidities. HIV–NCD comorbidity was defined as being HIV-positive and having at least one non-communicable disease. Chi-square tests and logistic regression analyses were performed to identify the determinants of HIV–NCD comorbidity. Among the 1,850 APLWH included in the analysis, 70.2% were female, 63.0% resided in rural areas, and 70.0% had attained primary education. Hypertension was the most prevalent chronic condition (3.3%), followed by lung disease (1.1%), cancer/tumor (1.0%), and heart disease (1.0%), respectively. The overall prevalence of HIV–NCD comorbidity was 6.7% (95% CI: 5.4–8.3). Female participants had significantly higher odds of HIV–NCD comorbidity (aOR = 1.9; 95% CI: 1.2–3.1; p = 0.010). Age was also strongly linked to comorbidity, with individuals aged 45–54 years (aOR = 9.4; 95% CI: 2.7–33.4; p = 0.001) and those aged ≥ 55 years (aOR = 16.1; 95% CI: 4.4–58.3; p < 0.001) showing higher odds than those aged 15–24 years. HIV–NCD comorbidities are common among APLWH. Female sex and older age significantly increase the risk, emphasizing the importance of tailored interventions for prevention, early detection, and integrated care strategies.

## Background

Human Immunodeficiency Virus (HIV) remains a significant global public health challenge despite decades of investment in prevention, diagnosis, and treatment. In 2024, an estimated 40.8 million people were living with HIV worldwide, with 97% of cases occurring among individuals aged ≥ 15 years (1). HIV-related morbidity and mortality continue to be substantial, with approximately 630,000 deaths globally attributed to HIV in 2024, 87% of which occurred in adults (1). Similarly, non-communicable diseases (NCDs), including cardiovascular diseases, cancers, diabetes, chronic respiratory conditions, and mental disorders, have become the leading causes of death worldwide (2–4). NCDs accounted for nearly 75% of all global deaths in 2021 (2), with low- and middle-income countries (LMICs) bearing 86% of this burden (2, 3, 5). This epidemiological transition has introduced complex healthcare challenges, particularly in settings already struggling with infectious diseases, such as HIV (6–11).

The increasing burden of non-communicable diseases among PLHIV has become a significant public health concern (3, 9, 12–16). While antiretroviral therapy (ART) has greatly improved survival, emerging evidence indicates that its long-term use, combined with chronic immune activation, contributes to an increased risk of metabolic complications and other NCD-related risk factors (6, 7).

As PLHIV live longer, comorbidities such as hypertension, diabetes, and cardiovascular diseases are more frequently emerging, potentially complicating HIV care and management (17). Therefore, the convergence of HIV and NCDs has created a dual disease burden, especially in sub-Saharan Africa, where both conditions are highly prevalent (18). Like other sub-Saharan African countries, Tanzania faces this challenge, with an HIV prevalence of approximately 4.4% in 2022 (19), along with a rising rate of NCDs driven by urbanization, changing lifestyles, socio-economic, demographic factors, and biological factors (18,20). The overlap between HIV and NCDs increases the vulnerability to poor health outcomes among people living with HIV (21). Therefore, understanding the burden and determinants of NCD comorbidities in people living with HIV is critical for integrated care models that address both infectious and chronic conditions (5, 22–24).

The Tanzania HIV Impact Survey (THIS) 2022-23 (19), which included questions on chronic diseases among participants, including those with HIV, provides a unique opportunity to assess the burden and determinants of NCD comorbidities among adults living with HIV in Tanzania. Thus, this study aimed to estimate the burden and identify the determinants of non-communicable disease comorbidities among adults living with HIV in Tanzania using data from the HIV Impact Survey.

## Material and Methods

### Ethics statement

Ethical approval for the original survey was granted by the institutional review boards (IRBs) of Columbia University Medical Centre, West, the CDC, the National Institute for Medical Research (NIMR), and the Zanzibar Health Research Institute (ZAHRI). This secondary analysis utilized publicly available anonymized data and was conducted in accordance with ethical guidelines to ensure the confidentiality of the participants.

### Data source and study design

The present analysis used data from the THIS 2022–2023, a nationally representative, cross-sectional, two-stage, population-based household survey of individuals aged 15 years or older. This survey was conducted between November 2022 and March 2023 and aimed to assess the effectiveness of Tanzania’s national and regional HIV response by providing estimates of HIV incidence, prevalence, and viral load suppression among adults living with HIV. It employed a robust multistage sampling strategy. A total of 39,442 adults aged 15 years or older were eligible to participate in all 31 regions of Mainland Tanzania and Zanzibar. Among them, 35,957 adults completed individual interviews, and 33,663 participants provided biomarker specimens (**Fig 1**). THIS 2022–2023 included home-based HIV testing with results returned to participants, along with counselling and linkage to antiretroviral therapy (ART) for those who tested positive and were not previously on treatment. For the current analysis, the study sample consisted of 1,850 individuals aged ≥ 15 years who tested HIV-positive (**Fig 1**).

**Fig 1:**
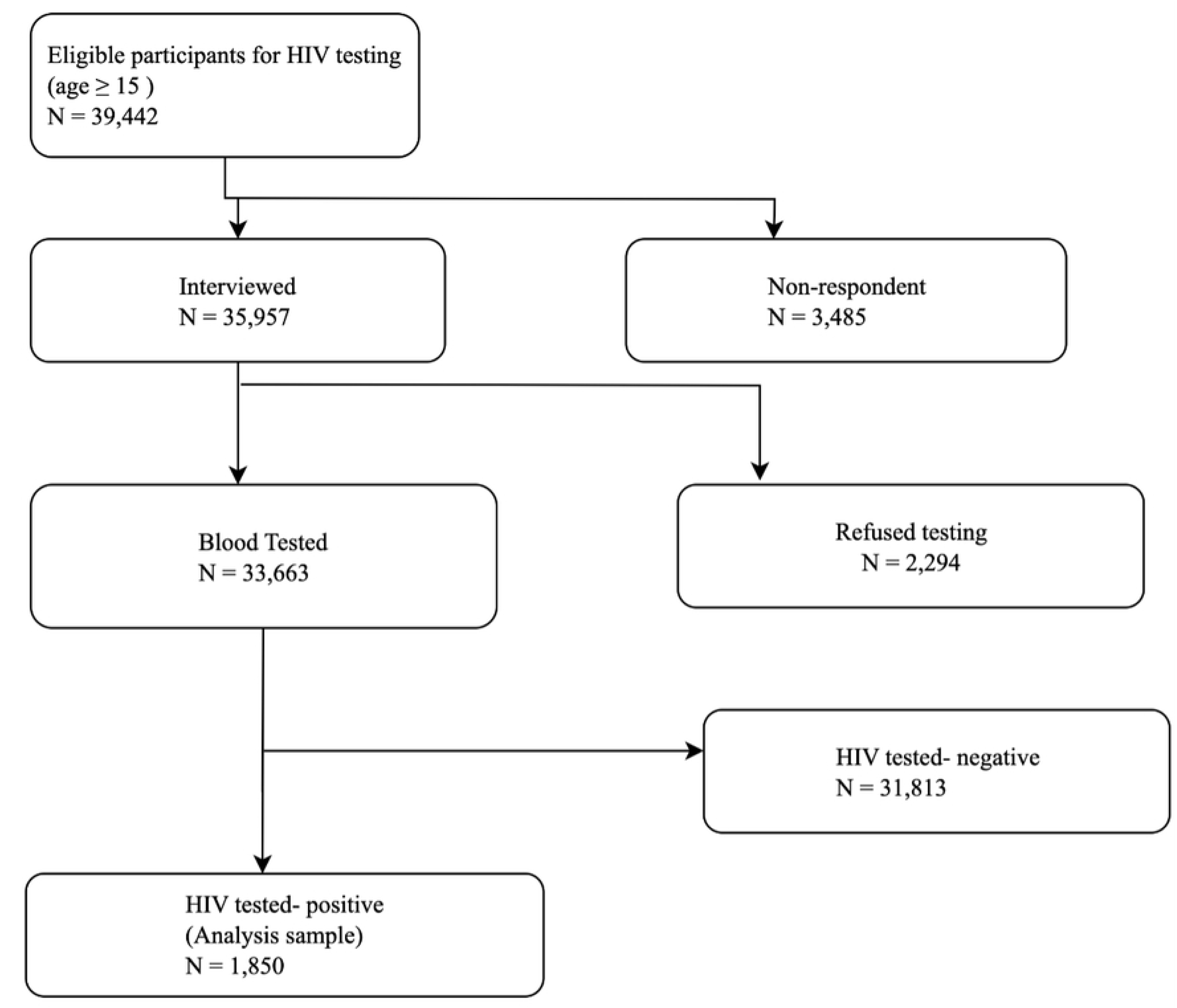
Flow diagram showing the study sample.

### Data extraction and quality assessment

Individual-level and biomarker datasets were extracted from the survey. As the data were pre-processed, cleaned, and recoded, we performed additional quality assurance checks to ensure data consistency. Initial exploratory analyses were conducted to identify any missing values or inconsistencies and verify the reliability of the dataset for subsequent analyses.

### Outcome and study variables

The outcome variable was HIV-NCD comorbidity, defined as HIV-positive status along with the presence of at least one non-communicable disease. Information on NCDs was collected through self-report, in which participants were asked whether a doctor or health worker had ever told them that they had a chronic health condition. The NCDs included in this definition were diabetes, hypertension, cardiovascular disease, kidney disease, cancer/tumour, chronic lung disease, and depression or other mental health disorders. HIV-NCD comorbidity was coded as a binary outcome (yes/no).

The predictor variables included sociodemographic and economic characteristics (age, sex, marital status, education level, place of residence, and wealth quintile), behavioral risk factors (smoking status and alcohol consumption), and clinical characteristics (ART use, WHO clinical stage, viral load suppression status, and ART regimen). These variables were analyzed as potential determinants of HIV–NCD comorbidities. Further details regarding the data source and study design are available elsewhere (19).

### Statistical Analysis

We performed descriptive analyses to summarise the distribution of study participants across socioeconomic, demographic, behavioural, and clinical characteristics (unweighted results). The results are presented in a table as frequencies and percentages. We then estimated the weighted prevalence of each non-communicable disease separately and in combination among people living with HIV. To assess associations between HIV-NCDs comorbidities and associated determinants, we performed cross-tabulations and applied the Chi-Square test. Furthermore, we conducted univariate (unadjusted) logistic regression analyses for each factor separately to examine its association with HIV-NCD comorbidity. Variables with a p-value < 0.2 in the unadjusted analyses were included in the multivariate (adjusted) logistic regression model to control for potential confounding. Variables with p<0.05 were considered determinants of HIV-NCD comorbidity.

To account for the complex sampling design, the data were weighted, and all analyses applied survey weights that incorporated clustering and stratification to ensure nationally representative estimates and valid standard error calculations. All analyses were performed using Stata version 17.

## Results

### Characteristics of study participants

The 2022–2023 Tanzania HIV Impact Survey included 33,663 individuals who underwent HIV testing, of whom 1,850 tested positive. The current analysis aimed to assess the burden and identify the determinants of non-communicable disease and HIV-NCD comorbidity among HIV-positive adults. The study sample comprised 1,850 participants, of whom 70.2% were female, and more than 63% resided in rural areas. The median age was 43.7(IQR:35-52), with the majority of participants aged between 35–54 years (56.1%). Over half (52.1%) were married or living together, and 17.4% were widowed. Approximately 70% of the participants had primary education, and 17% had no formal education. Behavioral risk factors were relatively uncommon, with 90.4% of participants being non-smokers and only 4.4% reporting alcohol use. Clinically, 80.8% had detectable ARVs, and 81.1% achieved viral load suppression of < 1000 copies/mL. WHO staging indicated that 56.5% were in stage 1, and advanced stages (3 and 4) accounted for 6.3%. Wealth distribution was relatively similar, although 24.4% were in the middle quintile. Geographically, most participants came from the Southern Highlands (28.2%) and Lake Zone (27.4%) (**Table 1**).

**Table 1:**
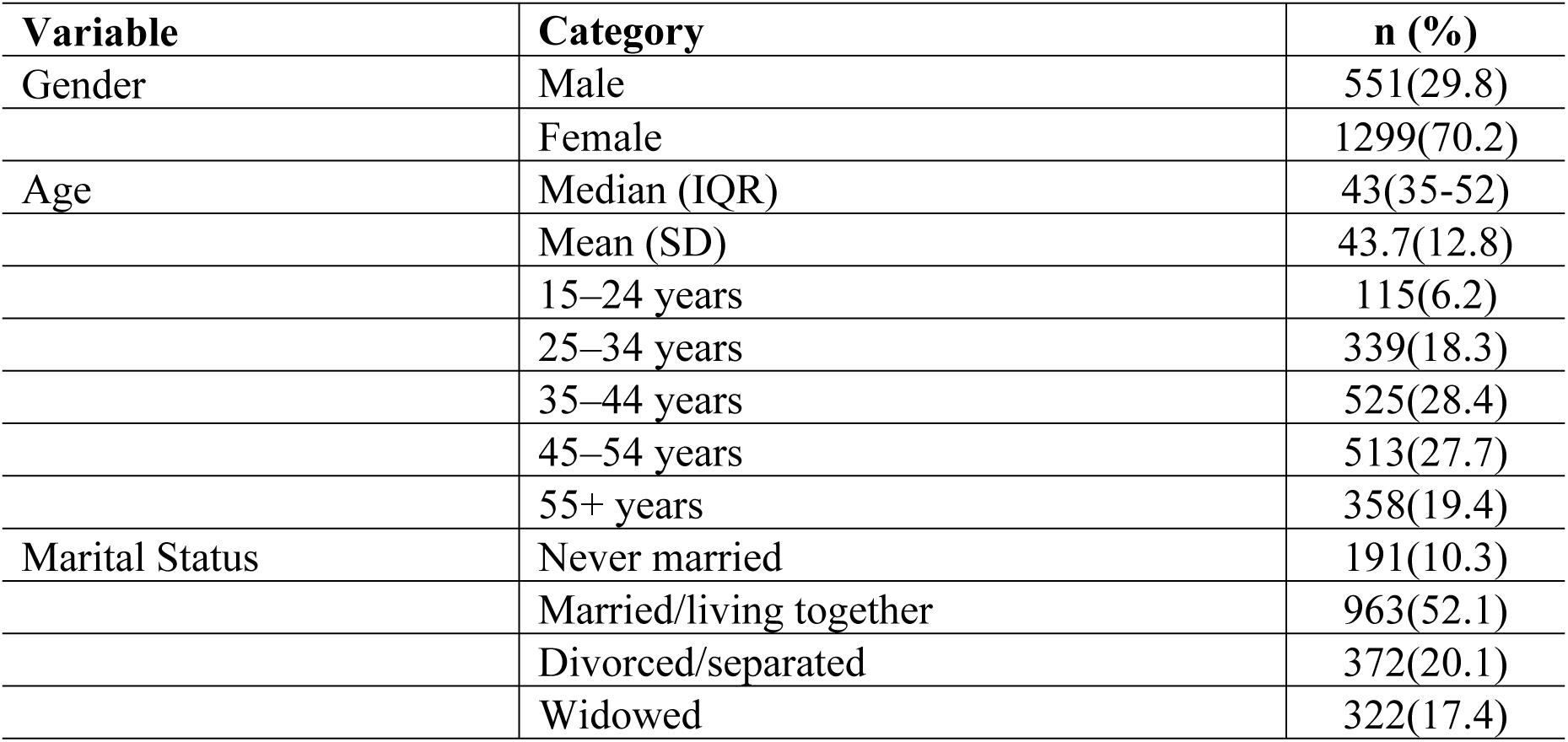
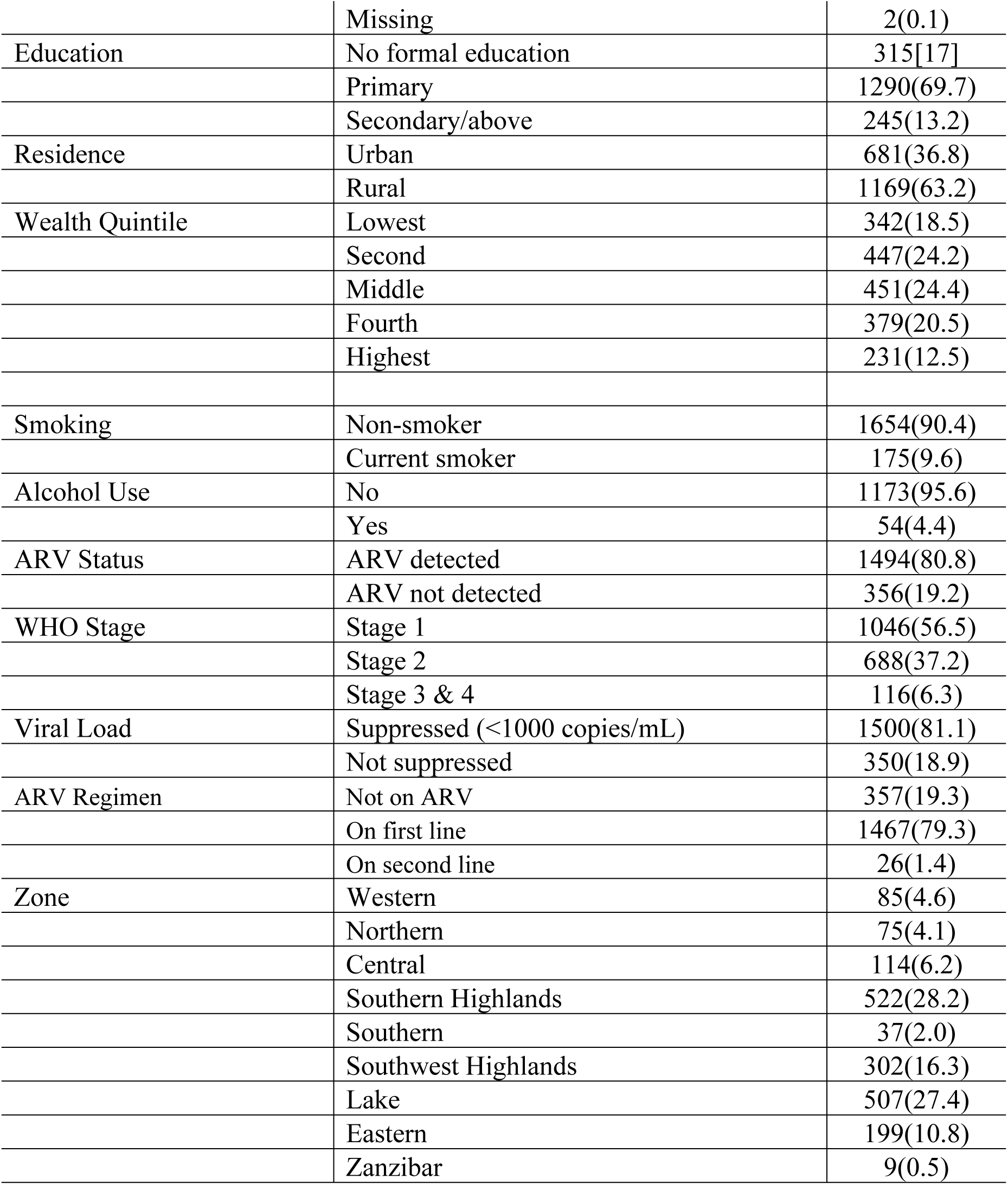
Demographic and clinical characteristics of people living with HIV (n = 1,850)

**Table 2:** Prevalence of self reported HIV-NCD comorbidities by demographic and clinical characteristics.

| Variable | Category | n | % with comorbidity (95%CI) | p-value |
| --- | --- | --- | --- | --- |
| Sex | Male | 551 | 5.0(3.4-7.3) | 0.048 |
|  | Female | 1299 | 7.5(6.0-9.4) |  |
| Age group | 15–24 yrs | 115 | 0.9(0.2-4.1) | <0.001 |
|  | 25–34 yrs | 339 | 1.7(0.6-4.3) |  |
|  | 35–44 yrs | 525 | 3.7(2.4-5.9) |  |
|  | 45–54 yrs | 513 | 9.5(7.1-12.6) |  |
|  | 55+ yrs | 358 | 15.0(10.6-20.7) |  |
| Marital status | Never married | 193 | 2.0(0.7-5.2) | 0.001 |
|  | Married | 963 | 6.1(4.5-8.2) |  |
|  | Divorced | 372 | 7.3(5.2-10.2) |  |
|  | Widowed | 322 | 11.5(7.8-16.6) |  |
| Education | No formal | 315 | 6.4(3.8-10.4) | 0.976 |
|  | Primary | 1290 | 6.8(5.2-8.7) |  |
|  | Secondary+ | 245 | 6.8(4.0-11.3) |  |
| Residence | Urban | 681 | 6.5(4.7-8.8) | 0.784 |
|  | Rural | 1169 | 6.9(5.1-9.1) |  |
| Wealth quintile | Lowest | 342 | 7.6(4.8-11.8) | 0.896 |
|  | Second | 447 | 5.6(3.2-9.6) |  |
|  | Middle | 451 | 6.6(4.4-9.7) |  |
|  | Fourth | 379 | 6.9(4.6-10.1) |  |
|  | Highest | 231 | 7.0(4.4-10.9) |  |
| Smoking | None | 1654 | 7.0(5.5-8.7) | 0.229 |
|  | Current | 175 | 4.3(2.0-9.2) |  |
| Alcohol use | No | 1173 | 6.6(5.2-8.4) | 0.453 |
|  | Yes | 54 | 10.0(3.1-27.9) |  |
| ARV detected | Yes | 1494 | 7.1(5.8-8.8) | 0.244 |
|  | No | 356 | 5.2(3.0-8.9) |  |
| WHO stage | Stage 1 | 1046 | 7.1(5.5-9.1) | 0.228 |
|  | Stage 2 | 688 | 5.5(4.0-7.6) |  |
|  | Stage 3 | 116 | 9.9(4.7-19.7) |  |
| Viral suppression | Yes | 1500 | 7.1(5.7-8.7) | 0.299 |
|  | No | 350 | 5.4(3.2-9.0) |  |
| ARV Regimen | Not on ARV | 357 | 5.2(3.02-8.81) | 0.344 |
|  | On first line | 1467 | 7.03(5.65-8.73) |  |
|  | On second line | 26 | 11.17(3.34-31-35) |  |
| Zone | Western | 85 | 5.1(0.8-27.8) | 0.895 |
|  | Northern | 75 | 6.4(3.2-12.4) |  |
|  | Central | 114 | 10.8(6.6-17.3) |  |
|  | Southern highlands | 522 | 8.2(5.2-12.7) |  |
|  | Southern | 37 | 13.3(7.0-23.7) |  |
|  | South west highlands | 302 | 6.5(4.3-9.8) |  |
|  | Lake | 507 | 5.5(4.0-7.7) |  |
|  | Eastern | 199 | 6.3(3.4-11.4) |  |
|  | Zanzibar | 9 | 0.0 |  |

### Prevalence of HIV-NCD comorbidities

The overall prevalence of comorbidities was 6.7% (95% CI: 5.4–8.3) (**Fig 2**). The prevalence increased with age, from 0.9% among young adults aged 15-24 years to 9.5% among those aged 45-54 years, reaching a peak of 15% in individuals aged 55 years and above. Additionally, the prevalence of each non-communicable disease among adults living with HIV was generally low across age groups (**Fig 2**). Hypertension was the most common chronic condition (3.3%) and increased with age, with the highest prevalence (8.3 %) observed among those aged ≥ 55 years. Other chronic conditions, such as lung disease (1.1%), cancer/tumor (1.0%), and heart disease (1.0%), were less common but also increased with age. Diabetes, kidney disease, and depression were less likely to be reported <1%), with the highest prevalence occurring in the oldest age group.

**Fig 2:**
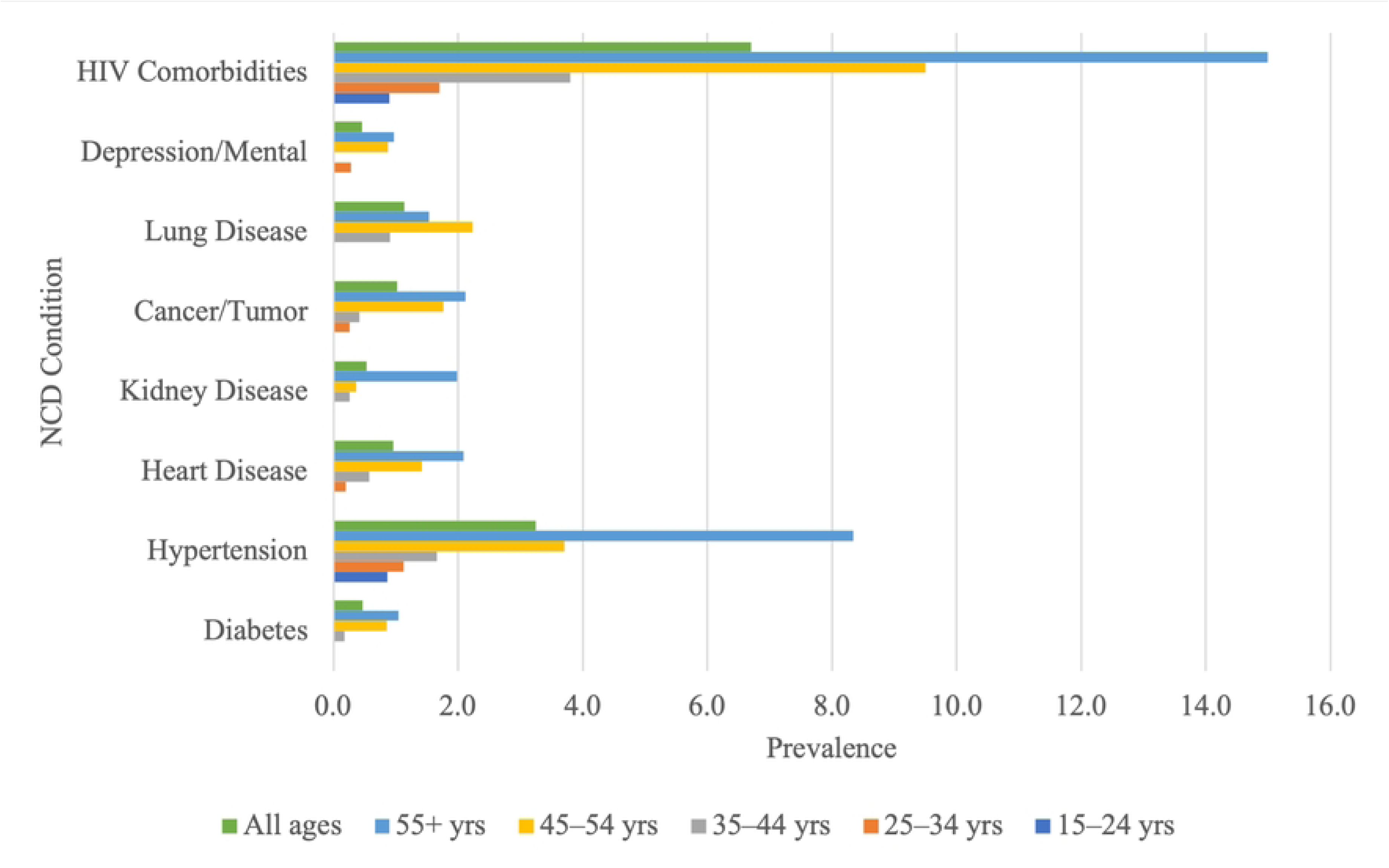
Prevalence of self-reported NCD and HIV comorbidities among people living with HIV disaggregated by age

### Association between comorbidities and demographic and clinical characteristics

Significant differences were observed by sex (p = 0.0475), with females showing a higher prevalence (7.5% [95% CI: 6.0-9.4%]) than males. The prevalence of HIV-NCD comorbidity varied significantly across age groups (p < 0.001) and increased from 0.9% in 15–24-year-olds to 15.0% in those aged ≥ 55 years. Marital status also showed significant variation (p <0.001), with widowed participants having the highest prevalence (11.5%) compared to those who had never married (2.0%). Other variables, including education, residence, smoking, alcohol use, ARV detection, WHO stage, viral load suppression, wealth status, and geographic zone, did not show significant differences in comorbidities across categories (p >0.05). However, the prevalence was high among alcohol users (10.0%) compared to non-users, 11.17% among those using second-line ARV regimen, 9.9% among those in WHO stage 3 and 4, and 13.3% among those living in the Southern zone. Notably, almost no individual had HIV-NCD comorbidity in Zanzibar.

### Determinants of HIV-NCD comorbidity

Of the 12 potential determinants of HIV-NCD comorbidities, only three–sex, age, and marital status–were initially found to be significantly (at p<0.2) associated with the prevalence of HIV-NCD comorbidities among people living with HIV (**Table 3**). Females had higher odds of having HIV-NCD comorbidities than males (uOR = 1.6, 95% CI: 1.0–2.4, p = 0.051). Age showed a strong positive association with HIV-NCD comorbidities with individuals aged 35–44 years (uOR = 4.4, 95% CI: 1.2–16.4, p = 0.027), 45–54 years (uOR = 12.0, 95% CI: 3.4–42.9, p < 0.001) and ≥ 55 years (uOR = 20.0, 95% CI: 5.5–73.1, p < 0.001) had significantly higher odds of HIV-NCD comorbidities than those aged 15–24 years. This pattern indicates a strong positive relationship between increasing age and the likelihood of HIV-NCD comorbidity. Regarding marital status, married or cohabiting individuals (uOR = 3.3, 95% CI: 1.2–8.6, p = 0.018), divorced or separated individuals (uOR = 4.0, 95% CI: 1.5–10.8, p = 0.009), and widowed individuals (uOR = 6.5, 95% CI: 2.4–17.9, p = 0.001) had significantly higher odds of HIV-NCD comorbidities than single participants. However, after adjustment in the weighted multivariable analysis, only sex and age remained significantly (p<0.05) associated with HIV-NCD comorbidities. Females had nearly two times higher odds of HIV-NCD comorbidities than males (aOR = 1.9, 95% CI: 1.2–3.1, p = 0.010). Age remained a strong predictor, with participants aged 45–54 years (aOR = 9.4, 95% CI: 2.7–33.4, p = 0.001) and those aged 55 years and above (aOR = 16.1, 95% CI: 4.4–58.3, p < 0.001) having significantly higher odds of HIV-NCD comorbidities than individuals aged 15–24 years.

**Table 3:** Determinants of HIV-NCD comorbidity among people living with HIV.

| Variable | Category | uOR(95%CI) | p-value | aOR(95%CI) | p-value |
| --- | --- | --- | --- | --- | --- |
| Gender | Male | Ref |  | Ref |  |
|  | Female | 1.6 (1.0–2.4) | 0.051 | 1.9 (1.2–3.1) | 0.010 |
| Age group | 15–24 | Ref |  | Ref |  |
|  | 25–34 | 1.93(0.4–10.5) | 0.432 | 1.5 (0.27–7.92) | 0.641 |
|  | 35–44 | 4.4 (1.2–16.4) | 0.027 | 3.3 (0.92–12.11) | 0.066 |
|  | 45–54 | 12.0 (3.4–42.9) | <0.001 | 9.4 (2.65–33.38) | 0.001 |
|  | 55+ | 20.03 (5.49–73.07) | <0.001 | 16.08 (4.44–58.25) | <0.001 |
| Marital status | Single | Ref |  |  |  |
|  | Married/cohabiting | 3.27 (1.24–8.59) | 0.018 | 2.09 (0.81–5.39) | 0.123 |
|  | Divorced/separated | 3.97 (1.46–10.80) | 0.009 | 2.32 (0.84–6.42) | 0.101 |
|  | Widowed | 6.51 (2.37–17.88) | 0.001 | 1.92 (0.69–5.33) | 0.198 |
| Education level | No formal education | Ref |  |  |  |
|  | Primary | 1.06 (0.60–1.90) | 0.828 |  |  |
|  | Secondary/above | 1.07 (0.48–2.36) | 0.866 |  |  |
| Place of residence | Urban | Ref |  |  |  |
|  | Rural | 1.06 (0.67–1.69) | 0.783 |  |  |
| Wealth status | Lowest quintile | Ref |  |  |  |
|  | Second | 0.73 (0.38–1.41) | 0.332 |  |  |
|  | Middle | 0.86 (0.42–1.73) | 0.653 |  |  |
|  | Fourth | 0.90 (0.49–1.64) | 0.719 |  |  |
|  | Highest | 0.92 (0.46–1.85) | 0.804 |  |  |
| Smoking status | None | Ref |  |  |  |
|  | Current | 0.60 (0.26–1.40) | 0.227 |  |  |
| Alcohol | No | Ref |  |  |  |
|  | Yes | 1.57 (0.36–6.95) | 0.536 |  |  |
| ARV use | Not on ARVs | Ref |  |  |  |
|  | On ARVs | 0.72 (0.40–1.28) | 0.251 |  |  |
| WHO Stage | Stage 1 | Ref |  |  |  |
|  | Stage 2 | 0.77 (0.49–1.18) | 0.219 |  |  |
|  | Stage 3&4 | 1.44 (0.67–3.08) | 0.333 |  |  |
| Viral Load | Suppressed (<1000 copies/mL) | Ref |  |  |  |
|  | Not suppressed | 0.75 (0.43–1.32) | 0.307 |  |  |
| ARV Regimen | Not on ARV | Ref |  |  |  |
|  | On first line | 1.38 (0.78–2.45) | 0.260 |  |  |
|  | On second line | 2.29 (0.43–12.25) | 0.318 |  |  |

## Discussion

This study provides critical insights into the burden and determinants of HIV-NCD comorbidities among adults living with HIV in Tanzania, using nationally representative data from the 2022–2023 HIV Impact Survey. Among the 1,850 HIV-positive adults analyzed, the overall prevalence of HIV-NCD comorbidities was 6.7%. However, hypertension was the most common condition and increased with age, reaching 8.3% among those aged ≥ 55 years. Other chronic conditions, such as lung disease, cancer, and heart disease, were less frequent but also increased with age, whereas conditions such as diabetes, kidney disease, and depression remained rare across all age groups. Age and sex were significant determinants of HIV-NCD comorbidity.

The higher burden of NCDs in older people living with HIV aligns with regional trends, suggesting a gradual but notable rise in HIV-NCD comorbidity, particularly as the HIV-positive population continues to age. Similar findings have been reported in a study conducted in Uganda, where HIV-NCD comorbidities, particularly hypertension, were the most common, with higher prevalence rates observed in older age groups (6). A comparable trend was observed in a study conducted in South Africa, which further reinforces the importance of age as a major determinant of comorbidity (7). Consistently, age emerged as the strongest predictor of HIV-NCD comorbidity in the present study, with adjusted odds ratios showing a sharp increase in risk among older adults, particularly those aged 45 years and above. Individuals in the 45–54 and 55+ age groups had 9.4 and 16.1times higher odds, respectively, of having NCD comorbidities than those aged 15–24 years. This pattern highlights the impact of aging as a key driver of the chronic disease burden in HIV-positive populations. This is consistent with findings from Ethiopia and Kenya, where older PLHIV were significantly more likely to develop hypertension and other NCDs (8,25).

Sex differences were also observed. Females accounted for 70% of the HIV-positive sample and had significantly higher odds of comorbidity than males. This may reflect a combination of factors, including biological vulnerability and gendered differences in health-seeking behavior, where more women are likely to seek health care services compared to men, making them aware of their health status (26,27). Moreover, available evidence indicates that women with HIV live longer than men (28). Longer survival with HIV may expose women to cumulative risks for chronic diseases, especially in the context of ART-related metabolic effects and immunological aging (29,30). This observation mirrors findings from a study in Rwanda, where women living with HIV were more likely to present with hypertension and other NCDs than men (31).

Notably, behavioral risk factors were not significantly associated with NCD comorbidities in this population. Smoking and alcohol consumption were relatively uncommon. This suggests that traditional lifestyle-related risk factors may not be the primary contributors to the observed comorbidities. Instead, non-modifiable factors, such as age and sex, along with HIV-related immunological changes, may play a more central role in driving the emergence of NCDs among people living with HIV. This finding is similar to a study conducted in Malawi, which found low levels of smoking and alcohol use among PLHIV and identified age as the dominant factor influencing the risk of NCDs (12). The low prevalence of smoking and alcohol consumption among PLHIV may be attributable to ongoing interventions, such as routine NCD screening and lifestyle counselling, implemented through Care and Treatment Clinics (CTCs)(32).

Marital status was significantly associated with comorbidities in the univariate analysis, with widowed individuals having the highest prevalence. However, this relationship lost statistical significance in the adjusted model, suggesting potential confounding factors such as age and sex. This finding contrasts with a study conducted in Northwestern Tanzania and Southern Uganda, where marital status remained a significant determinant of comorbidity even after adjustment, possibly reflecting different social support or access to care dynamics (18).

Other factors, such as education level, place of residence (urban vs. rural), wealth quintile, ARV status, WHO clinical stage, and viral suppression status, did not show significant associations with comorbidity, highlighting that HIV-related clinical indicators may not be the strongest determinants of NCD risk in this population. This aligns with previous studies indicating that as ART access and viral suppression improve across sub-Saharan Africa, traditional clinical markers may become less predictive of broader health outcomes, including NCDs (33).

From a public health perspective, there is a burden of NCDs in this population group. The strong association with age and gender signals an emerging challenge as Tanzania’s HIV-positive population continues to age and survive longer on ART regimens. Therefore, early integration of NCD screening and prevention into routine HIV care is warranted, particularly targeting older adults and women. Such integration could improve long-term outcomes, reduce the dual disease burden, and ensure the sustainability of HIV treatment programs as the epidemiological profile of PLHIV continues to evolve. Similar calls for integration have been emphasized in WHO regional guidelines and in studies across sub-Saharan Africa advocating for chronic disease management within HIV platforms (5,12,24).

This study had several limitations. First, the cross-sectional design limits causal inference between the determinants and comorbidities. Second, self-reported data for some NCDs may have led to an underestimation of their true prevalence in this study and may also be affected by individual health seeking behaviors and service coverage and availability patterns. Third, the low prevalence of some NCDs may have reduced the statistical power to detect significant associations. Moreover, the analysis was limited to individuals aged ≥ 15 years and considered a limited range of sociodemographic factors, without incorporating lifestyle factors which strongly influence NCD risk. The exclusion of these variables limits the ability of the study to capture the full spectrum of determinants of NCDs among people living with HIV. Nonetheless, the use of nationally representative data strengthens the generalizability of these findings. Additionally, this study provided important information for understanding the burden of NCDs in PLHIV in Tanzania and identified determinants, providing a foundation for monitoring and targeting interventions in regions with increased risk.

## Conclusion

Our study found that the burden of non-communicable disease comorbidities was relatively high among people living with HIV in Tanzania. Being female and over 45 years of age significantly increased the likelihood of having both HIV and NCDs. This highlights the urgent need for sustainable integrated healthcare services that address both HIV and NCDs, focusing on prevention, early detection, and management to improve the health outcomes of individuals living with HIV.

## Author contributions

Conceptualization, CBK, HRS, AML, GK, MTM, and JMM; methodology, AML, JJM, GPM and GK; software, AML, JJM, GPM and GK; formal analysis, AML, JJM, GPM and GK; data curation, CBK, HRS, AML, and GK.; writing - original draft, CBK, HRS, AML, and GK.; writing - review and editing, CBK, HRS, AML, GK, MTM, JJM, PMH, GPM, IRM, PM, SEM, and JMM; visualization, GK; supervision, MTM and JMM.; Resources, MTM and JMM. All authors have read and approved the final manuscript.

## Acknowledgments

We acknowledge PEPFAR and the U.S. Centers for Disease Control and Prevention (CDC) for their financial support through the National Institute for Medical Research (NIMR-Tanzania) under Cooperative Agreement Number NU2GGH002283. The views expressed in this paper are solely those of the authors and do not necessarily reflect the official positions of the CDC or NIMR.

## Data availability statement

The datasets used in this study are available online and can be accessed through https://phia-data.icap.columbia.edu/datasets upon submitting a request.

